# Latent clinical-anatomical dimensions of schizophrenia

**DOI:** 10.1101/2020.03.25.20040592

**Authors:** Matthias Kirschner, Golia Shafiei, Ross D. Markello, Carolina Makowski, Alexandra Talpalaru, Benazir Hodzic-Santor, Gabriel A. Devenyi, Casey Paquola, Boris C. Bernhardt, Martin Lepage, M. Mallar Chakravarty, Alain Dagher, Bratislav Misic

## Abstract

Widespread structural brain abnormalities have been consistently reported in schizophrenia, but their relation to the heterogeneous clinical manifestations remains unknown. In particular, it is unclear whether anatomical abnormalities in discrete regions give rise to discrete symptoms, or whether distributed abnormalities give rise to the broad clinical profile associated with schizophrenia. Here we apply a multivariate data-driven approach to investigate covariance patterns between multiple symptom domains and distributed brain abnormalities in schizophrenia. Structural MRI, and clinical data were derived from one discovery sample (133 patients, 113 controls) and one independent validation sample (108 patients, 69 controls). Disease-related voxel-wise brain abnormalities were estimated using deformation based morphometry. Partial least squares analysis was used to comprehensively map clinical, neuropsychological and demographic data onto distributed deformation in a single multivariate model. The analysis identified three latent clinical-anatomical dimensions that collectively accounted for 55% of the covariance between clinical data and brain deformation. The first latent clinical-anatomical dimension was replicated in an independent sample, encompassing cognitive impairments, negative symptom severity and brain abnormalities within the default mode and visual networks. This cognitive-negative dimension was associated with low socioeconomic status and was represented across multiple races. Altogether, we identified a continuous cognitive-negative dimension of schizophrenia, centered on two intrinsic networks. By simultaneously taking into account both clinical manifestations and neuroanatomical abnormalities, the present results open new avenues for multi-omic stratification and biotyping of individuals with schizophrenia.

## INTRODUCTION

Schizophrenia is characterized by heterogeneous clinical manifestations including positive symptoms, negative symptoms and generalized cognitive impairments. This complex clinical pattern is already prevalent prior to and during first-episode psychosis^1^. While positive symptoms tend to reduce over time, negative and cognitive symptoms are more likely to persist over time, severely affecting long-term social functioning and quality of life^2–9^.

Convergent findings from neuroimaging link clinical manifestations of schizophrenia with widespread disruption of structural and functional brain networks^10–13^. Several large scale studies and meta-analyses provide evidence for widespread anatomical alterations, including reduced cortical thickness, subcortical volume and white matter integrity^14–16^. These localized brain abnormalities have individually been linked to clinical manifestations of positive, negative and cognitive symptoms^17–20^.

But how do complex clinical phenotypes map onto distributed brain networks? The organization of brain connectivity increases the likelihood that local pathological perturbations affect synaptically-connected neuronal populations^21^. Thus, structural abnormalities with a distributed topography may reflect the underlying network architecture and manifest as a diverse set of cognitive and affective symptoms^22–25^. Recent studies have demonstrated such links between brain structure and function both in healthy controls^26,27^, and across a number of neurological and psychiatric diseases^28–32^.

Several methodological limitations might have hampered the progress to identify comprehensive clinical-anatomical signatures of schizophrenia. First, the heterogeneity of clinical manifestation cannot be captured by case-control designs, or studies focusing on a single symptom domain (e.g. only positive or only negative symptoms). Second, many previous studies were designed to capture associations between symptom dimensions and global brain measures or localized brain changes with *a priori* defined regions of interest. Altogether, previous work eschews the possibility of a pleiotropic-like mapping between anatomy and function, whereby distributed structural alterations may simultaneously lead to multiple positive and negative symptoms^17–20,33^.

The relationship between anatomical abnormalities and clinical manifestation is particularly important for understanding heterogeneity in the patient population. Recent efforts have been directed towards stratifying individuals into non-overlapping clusters or biotypes based either on clinical-behavioral features^34^ or neuroimaging features^35^. Although promising, such “hard partitioning” methods are designed for precise categorical stratification based either on clinical/behavorial or neuroimaging measures, but do not consider the possibility of continuous phenotypic dimensions that span multiple clinical domains, nor do they explicitly integrate clinical and neuroanatomical features. By focusing on single “modalities” (clinical only or imaging only), unsupervised learning methods miss out on the critical link between brain and behavior, and may yield solutions that are difficult to interpret or reconcile with clinical experience^30^. Thus, identifying continuous clinical-anatomical dimensions would complement categorical biotyping efforts,helping to situate individuals and biotypes in a wider multivariate space defined by both clinical presentation and anatomical abnormalities^36,37^.

Here we apply a data-driven method to identify multi-modal phenotypic axes of schizophrenia. Specifically, we use multivariate mapping between whole brain anatomical alterations and clinical symptoms in schizophrenia to reveal latent clinical-anatomical dimensions. We first estimate grey matter abnormalities in a sample of *N* =133 individuals with chronic schizophrenia and *N* = 113 healthy controls from the Northwestern University Schizophrenia Data and Software Tool (NUSDAST; http://schizconnect.org)^38^. Deformation-based morphometry (DBM) was applied to T1-weighted MR images to estimate cortical and subcortical grey matter tissue volume loss in patients with schizophrenia relative to healthy controls (hereafter referred to as “deformation”)^39–44^. We then identify disease-related deformation patterns using partial least squares analysis (PLS) (Fig. 1)^45–47^. The technique isolates patterns of deformation directly related to multiple symptom dimensions (including positive, negative and cognitive symptoms) and demographic data (Table I). We first validate the results in an independently collected dataset. We then investigate whether the spatial patterning of deformation is related to the intrinsic functional architecture of the brain. Finally, we link the most reliable clinical-anatomical dimensions to broader societal variables of interest, including socioeconomic status and race.

**Figure 1.**
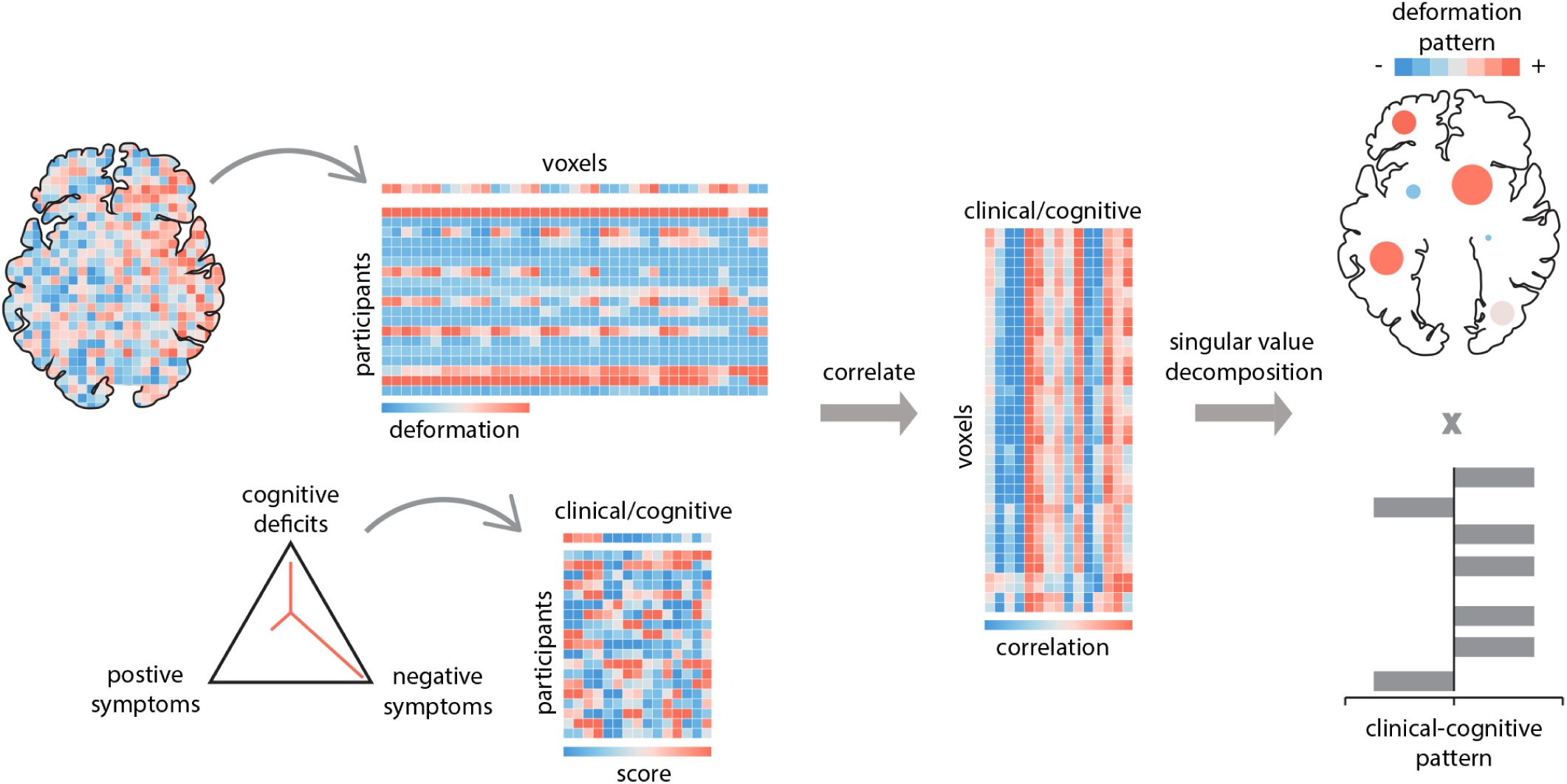
Partial least-squares analysis. PLS is a form of reduced-rank regression used to relate two sets of variables to each other. The original variables are correlated across participants and subjected to singular value decomposition. The decomposition yields multiple latent variables: linear combinations of the original variables, with the weights chosen to maximize the covariance between them. The contribution of individual variables to the latent variable is assessed by bootstrap resampling. The pairing of the deformation and clinical-cognitive pattern is assessed by permutation tests and cross-validation.

**TABLE 1.** Sample characteristics. Clinical, behavioral and demographic characteristics. (**A**) Discovery sample (NUSDAST). ^*a*^ based on 131 patients; ^*b*^ based on 86 patients; ^*c*^ based on 124 patients. (**B**) Validation sample (Douglas Institute). ^*d*^ Normalized composite cognitive score estimated from the CogState Research Battery protocol^102^ that includes cognitive domains of verbal memory, visual memory, working memory, processing speed, executive function, visual attention, and social cognition.*<.01, **<.001

| **(A) Discovery data (NUSDAST)** | **SCZ (n=133)** | **HC (=113)** | **Statistic** |
| --- | --- | --- | --- |
| age, mean (sd) | 34.8(13.2) | 23.5(8.4) | t=7.85** |
| female sex, no. (%) | 48(36.1) | 64(56.6) | x2=10.4* |
| years in school, mean (sd) | 12.2(2.3) |  |  |
| age of onset, mean (SD)<sup>a</sup> | 21.6(8.0) |  |  |
| years illness duration, mean (SD)<sup>a</sup> | 12.9(12.5) |  |  |
| SES score, mean (SD) | 45(15.5) |  |  |
| WMS cognitive, mean (SD) | -0.00005(4.9) |  |  |
| WAIS score, mean (SD) |  |  |  |
| matrix | 8.9(3.3) |  |  |
| vocabulary | 7.9(3.6) |  |  |
| ESRS score, mean (SD) |  |  |  |
| global | 1.2(1.9) |  |  |
| total | 5.5(6.6) |  |  |
| SANS score, mean (SD) |  |  |  |
| avolition/apathy | 10.7(6.9) |  |  |
| avolition/apathy - global | 4.3(2.4) |  |  |
| diminished expression | 10.3(9.5) |  |  |
| diminished expression - global | 3.0(2.2) |  |  |
| SAPS score, mean (SD) |  |  |  |
| disorganization | 5.1(5.8) |  |  |
| disorganization - global | 1.8(1.7) |  |  |
| reality distortion | 13.5(12.7) |  |  |
| reality distortion - global | 3.2(2.5) |  |  |
| chlorpromazine equivalent dosage, mean (SD)<sup>b</sup> | 391.3(379.8) |  |  |
| information on medication status obtained, no. (%)<sup>c</sup> | 93.2 |  |  |
| type of medication<sup>c</sup>, % |  |  |  |
| typical | 12.9 |  |  |
| atypical | 70.2 |  |  |
| both | 12.2 |  |  |
| unmedicated | 4.8 |  |  |
| **(B) Validation data (Douglas Institute)** | **SCZ (n=108)** | **HC (n=69)** | **Statistic** |
| age, mean (sd) | 35.2(8.2) | 34.1(9.0) | t=0.84 |
| female sex, no. (%) | 26(24.0) | 21(30.4) | x2=0.87 |
| WASI full-scale IQ, mean (SD) | 95.23(14.58) |  |  |
| Cognitive composite score, mean (SD)<sup>d</sup> | -5.5(5.6) |  |  |
| SANS score, mean (SD) |  |  |  |
| avolition/apathy | 12.3(7) |  |  |
| avolition/apathy - global | 5.8(2.1) |  |  |
| diminished expression | 10.3(7.1) |  |  |
| diminished expression - global | 3.7(2.0) |  |  |
| SAPS score, mean (SD) |  |  |  |
| disorganization | 5.6(6.5) |  |  |
| disorganization - global | 2.4(2.1) |  |  |
| reality distortion | 12.8(13.7) |  |  |
| reality distortion - global | 4.2(3.1) |  |  |
| chlorpromazine equivalent dosage, mean (SD) | 798.3(825.3) |  |  |

## METHODS

### Discovery dataset: NUSDAST

The discovery dataset was derived from the North-western University Schizophrenia Data and Software Tool (NUSDAST)^38^, downloaded from XNAT Central (http://central.xnat.org/) and the SchizConnect data sharing portal (http://schizconnect.org/). Briefly, the NUSDAST dataset is a cohort of individuals with schizophrenia, their non-psychotic siblings, healthy controls and their siblings. Detailed information is available at^38^. The final dataset used in this study comprised 133 individuals with schizophrenia and 113 healthy controls. Detailed inclusion criteria are included in the *Supplementary Methods*, while the selection flowchart is shown in Fig. S1.

### NUSDAST clinical and demographic data

Clinical and demographic data were derived from the baseline visit provided by the NUSDAST database (Table I). Among the demographic measures, we used age, sex/gender, years of schooling, and socioeconomic status (SES). The clinical assessment included the Scale for the Assessment of Positive Symptoms (SAPS;^48^) and the Scale for the Assessment of Negative Symptoms (SANS;^49^). Following the four symptom dimension approach from Kotov and colleagues^50^ as well as Strauss and colleagues (for two negative symptom factors, respectively)^51^, we calculated two negative symptom factors and two positive symptom factors. The two negative symptom dimensions comprised the SANS diminished expression factor (including affective flattening, alogia), and the SANS Avolition-Apathy factor (including avolition and anhedonia). The two positive symptom dimensions comprised the SAPS Reality distortion factor (including hallucinations and delusions) and the SAPS Disorganization factor (including bizarre behavior and thought disorder). These four factors were calculated for individual items (Sum Scores) and global ratings separately, resulting in a total of eight factors. Furthermore, total and global scores of the Extrapyramidal Symptom Rating Scale (ESRS) were included to assess four types of drug-induced movement disorders (DIMD) caused by antipsychotic treatment: parkinsonism, akathisia, dystonia, and tardive dyskinesia^52,53^.

Overall cognitive functioning was assessed with a composite score following the method suggested by Czepielewski and colleagues^18^. The composite score (WMS Cog) included the sum of the z-transformed scores on Logical Memory, Family Pictures, Letter-Number Sequencing, Spatial Span, and Digit Span from the Wechsler Memory Scale (WMS-III;^54^). Finally, individual scaled scores from the WAIS-III Matrix Reasoning and Vocabulary subsets were included as measures of executive function^38^ and crystallized knowledge (i.e., premorbid crystallized intellectual functioning (ePMC-IQ)^18^), respectively. Altogether, 15 demographic and clinical measures were entered in the PLS analysis to identify latent clinical-anatomical dimensions related to multiple symptom dimensions. SES, age of onset, duration of illness and antipsychotic medication (chlorpromazine equivalents) were left out from the PLS analysis and their relation with the final statistical model (clinical-anatomical dimensions) was tested post-hoc (for details see *Supplementary Methods*).

### NUSDAST neuroimaging data

All MRI scans were acquired on the same 1.5 T Vision scanner platform (Siemens Medical Systems) at the Mallinckrodt Institute of Radiology at Washington University School of Medicine^38^. Automated pre-processing was performed using the `minc-bpipe-library` pipeline (https://github.com/CobraLab/minc-bpipe-library) following manual quality control to remove scans with insufficient quality, see *Supplementary Methods*. Local change in the brain tissue’s volume density was calculated using Deformation-Based Morphometry (DBM;^55^). We interpret regional DBM values as measures of tissue loss or tissue expansion^39–43^. Note however that morphometric techniques do not directly measure the underlying cellular morphology and constitute a statistical model of physiological changes. DBM is estimated based on the deformation applied at each voxel to non-linearly register each brain to a given template. For details of the DBM pipeline, please see *Supplementary Methods*. Chronological age was regressed from DBM values prior to PLS analysis in both the NUSDAST and Douglas datasets.

### Validation dataset: Douglas Institute

T1-weighted MRI scans of 108 individuals with schizophrenia and 69 healthy controls were obtained from an independently collected dataset to validate the original findings (Table I). Details about the participant inclusion criteria, MRI acquisition, and data pre-processing are available elsewhere^56^ and also described in the *Supplementary Information*. Regional DBM values and clinical/cognitive measures overlapping with the discovery set were used for further analysis.

### Partial least squares

We used partial least squares (PLS) analysis to investigate the relationship between local changes in deformation (DBM values) and clinical/behavioral measures (Fig. 1). PLS analysis is a multivariate statistical technique that identifies weighted patterns of variables in two given sets or data blocks that maximally covary with each other^45–47^. In the present analysis, one variable set corresponded to deformation and the other to clinical measures. The two variable sets were correlated with each other across patients, and the resulting correlation matrix was subjected to singular value decomposition to identify latent clinical-anatomical dimensions.

Inference and validation of the statistical model was performed using nonparametric methods: (a) statistical significance of overall patterns was assessed by permutation tests^57^ ; (b) feature (voxel, clinical measure) importance was assessed by bootstrap resampling^58^ ;out-of-sample correlations between projected scores were assessed by cross-validation^59^ ; (d) stability of deformation and clinical patterns was assessed by split-half resampling^60^. Mathematical details of the analysis and inferential methods are described in *Supplementary Methods and Results*.

## RESULTS

### Clinical-anatomical dimensions of schizophrenia

Multivariate PLS analysis identified three statistically significant latent variables (LVs) that represent pairings between distributed deformation patterns (estimated by age-corrected DBM) and clinical-behavioral measures (Fig. 2a; LV-1: permuted *P* = 7.3 ×10^−3^; LV-2: permuted *P* = 5 ×10^−4^; LV-3: *P* = 7 ×10^−4^). These patterns respectively account for 27.5, 15, and 13% (total of 55.5%) of the covariance between clinical-behavioral data and brain deformation. Based on effect size and reliability (see below), we focus on LV-1 in the main text.

**Figure 2.**
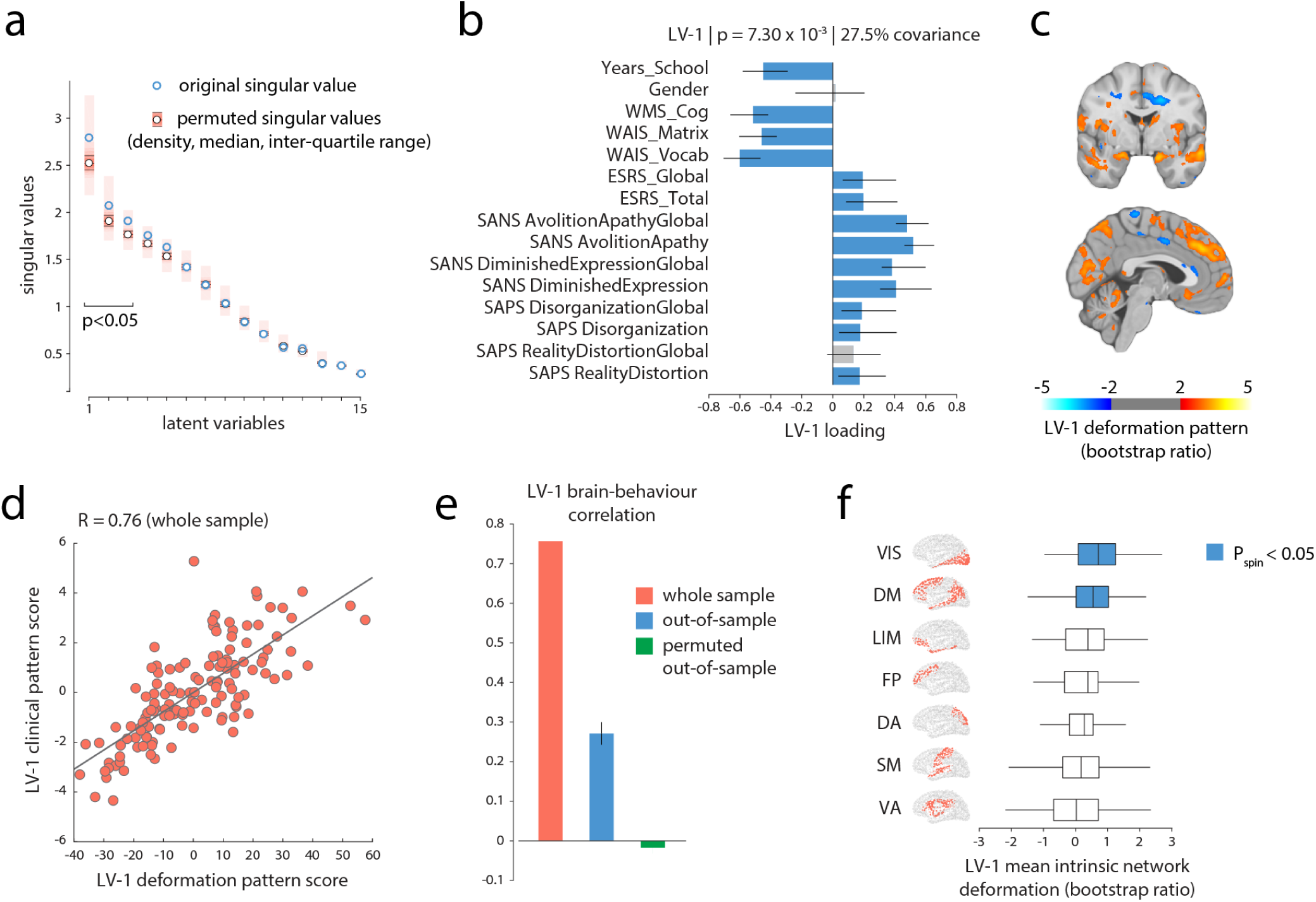
A clinical-anatomical signature of schizophrenia. (a) Partial least-squares analysis detected three statistically significant latent variables, mapping distributed patterns of deformation to clinical-behavioral characteristics. The first latent variable (LV-1) accounted for 27.5% of the covariance between the MRI and clinical-behavioral data. (b) Clinical features of LV-1. The contribution of individual clinical measures is shown using correlations between patient-specific clinical scores and scores on the multivariate pattern (loadings). Error bars indicate bootstrap-estimated standard errors. (c) LV-1 deformation pattern. The contribution of individual voxels is shown using bootstrap ratios (ratios between voxel weights and bootstrap-estimated standard errors, interpretable as z-scores; see *Supplementary Methods* for more detail). The deformation pattern is displayed on an MNI template (MNI152_symm_2009a; (*x* = −3, *y* = −2)). Patients who display this deformation pattern tend to score higher on measures of clinical severity of negative symptoms (e.g. SANS Avolition-Apathy) and tend to score lower on cognitive measures (e.g. WAIS). (d) Individual patient data is projected onto the weighted patterns shown in (b) and (c) to estimate scalar patient scores that quantify the extent to which individual patients express each pattern in LV-1. The two scores are correlated, suggesting that patients who display the deformation pattern in (c) tend to express the clinical phenotype in (b). (e) Correlations between deformation and clinical scores in the original sample (red; same as panel e), in held-out data (blue) and in a permuted null (green). (f) Specific intrinsic-network deformation. The PLS-derived deformation pattern is stratified into resting-state networks (RSNs) defined by Yeo and colleagues^61^. The bars indicate mean deformations for each network. *P*-values are estimated with respect to the spin test null developed by Alexander-Bloch and colleagues^62^. Yeo networks: DM = default mode, DA = dorsal attention, VIS = visual, SM = somatomotor, LIM = limbic, VA = ventral attention, FP = fronto-parietal.

Fig. 2b shows the loadings (i.e. correlations) of individual clinical and cognitive scales with the first latent variable (LV-1). The strongest contributors to LV-1 were cognitive deficits (all *r <* −.45), severity of negative symptoms (all *r >* .38) and educational attainment (*r* = −.45). Positive symptoms and DIMD also contributed to LV-1 but to a lesser extent (all *r >* .15 but *<* .2). In other words, LV-1 captures predominantly clinical features of the cognitive and negative symptom domains (cognitive-negative dimension).

Fig. 2c shows the corresponding deformation pattern associated with LV-1, indexed by bootstrap ratios. Briefly, bootstrap ratios measure the reliability of each weight across participants, and can be interpreted as a z-score (see *Supplementary Methods* for more detail). This brain deformation pattern is comprised of occipital (visual), medial parietal, lateral temporal, prefrontal (medial pre-frontal cortex, superior frontal gyrus), limbic and paralimbic regions including the cingulate (anterior and posterior) and hippocampus. In addition, the deformation pattern involves subcortical regions including the caudate and cerebellar structures. Altogether, the first latent clinical-anatomical dimension indicates that distributed deformation in this distributed network of regions is associated with negative symptom severity and lower cognitive performance. Finally, clinical and deformation scores were then correlated (Fig. 2d); the mean out-of-sample correlations were *r* = 0.27 (Fig. 2e). Further details on cross-validation in *Supplementary Results*.

LV-2 and LV-3 are shown and described in detail in the *Supplementary Results* (Fig. S2b,c). Associations between patient-specific scores of all three latent variables and age of onset, duration of illness, and medication dosage are reported in *Supplementary Results*.

### External replication

To further assess the reliability of the results, we validated the PLS-derived patterns in an independently acquired replication dataset (Douglas dataset; *N* = 108 individuals with schizophrenia; see *Methods*). Regional DBM values from the validation set (Douglas) were projected onto the PLS model derived from the discovery set (NUSDAST) to estimate the predicted brain deformation scores for the validation set. The predicted brain deformation scores were then correlated with the 12 clinical, cognitive and demographic measures that were common to two datasets, yielding a predicted clinical profile for the validation set (Fig. S3, left column). The discovery and validation clinical profiles were then correlated and the significance of correlations were tested against a permuted null model (1,000 repetitions; Fig. S3, middle column). Finally, bootstrap resampling was used to generate a distribution of correlations between the discovery and validation profiles (1,000 repetitions; Fig. S3, right column).

For the first latent variable (cognitive-negative dimension), we find a significant association between the clinical profiles of the discovery and validation datasets (*r* =0.6, *P* = 2.0 *×* 10^−2^, (95% CI: [0.09 0.90]) (Fig. S3). In other words, projecting the brain deformation data from the validation set on the first latent variable of the discovery revealed a similar cognitive-negative clinical profile with 36% of variance explained. Thus, we were able to partly replicate the clinical-anatomical dimension of the first latent variable in an independent validation dataset. Repeating the same analysis for LV-3 revealed a positive but non-significant association between the clinical profiles of the discovery and validation datasets (*r* = 0.42,*P* = 1.09 *×*10^−1^, (95% CI: [−0.60 0.92]) and no significant association between the clinical profiles of LV-2(*r* = −0.50, *P* = 5.8 × 10^−2^, (95% CI: [−0.87 0.26]). Please note that the discovery (NUSDAST) and replication dataset (Douglas) differed significantly in several aspects including ethnicity (NUSDAST: mixed Caucasian and African-American, Douglas: Caucasian), fewer female participants (*χ*_2_ = 4.04, *P* = 4.4 × 10^−2^), higher antipsychotic medication (*t* = 4.49, *P <* 1.0 × 10^−*4*^) and higher global positive and negative symptoms in the discovery sample (SAPS Disorganization Global, *t* = 2.38, *P* = 1.8*×* 10^−2^; SAPS RealityDistortion Global, *t* = 2.55, *P* = 1.1*×* 10^−2^; SANS Avolition-Apathy Global, *t* = 2.75, *P* = 6.0 *×*10^−3^; SANS Diminished Expression Global, *t* = 2.55, *P* = 1.1 × 10^−2^). Although these marked differences might have hampered replication of all three clinical-anatomical dimensions, the most prominent clinical profile of the first latent variable is still represented in the independent replication dataset.

### Clinical-anatomical dimensions map on intrinsic networks

We next asked how clinically defined deformation patterns are topographically distributed in the brain, and whether their organization reflects the underlying functional architecture. The deformation pattern corresponding to the clinical features of LV-1 (cognitive-negative dimension) appears to mainly target brain regions associated with the default mode network and visual network (Fig. 2c). To statistically assess if this is the case, we used a recently-developed spatial permutation procedure^62^. We stratified voxels according to their membership in seven intrinsic networks and calculated the mean bootstrap ratio value within each network^61^. To construct a null distribution for network means, we projected the data on a sphere and randomly rotated the sphere, permuting the intrinsic network labels of brain regions but preserving the spatial autocorrelation of the map^62,63^. The mean bootstrap ratio was then re-calculated for each network for the permuted sample. The procedure was repeated 10,000 times to construct a distribution of network means under the null hypothesis that regional volume loss patterns are independent of affiliation with specific intrinsic networks.

Fig. 2e shows the mean bootstrap ratios for each network. Consistent with the voxel-wise anatomical pattern in Fig. 2c, deformation of the cognitive-negative dimension (LV-1) was significantly greater in the default mode and visual networks than expected by chance (*P* = 1.2 ×10^−2^ and *P* = 3.5 ×10^−2^, respectively), demonstrating a specific spatial mapping within these two intrinsic networks. Complete results for network specificity of deformation patterns of LV-2 and LV-3 are presented in the *Supplementary Results* and Fig. S2 (right column).

### Clinical-anatomical dimensions are associated with lower socioeconomic status

In schizophrenia, socioeconomic status (SES) is a predictor for increased risk of hospitalization^64,65^, symptom severity^66,67^, poor outcome^68,69^ and has been shown to be associated with brain function and structure^70,71^. Using simple correlations, we investigated the relation between the clinical features and corresponding deformation patterns of the first clinical-anatomical dimension with SES. We observed that both brain deformation and clinical features of LV-1 were associated with SES (anatomical: *r* = 0.36, *P* = 1.8×10^−05^, 95% CI [.20,.50], clinical: *r* = 0.28, *P* = 1.0 × 10^−03^, 95% CI [.11,.43]). To illustrate this association, we colored the individual points, corresponding to patients, according to their SES (Fig. 3a). Taken together, the clinical-anatomical signature of the cognitive-negative dimension is associated with lower SES.

**Figure 3.**
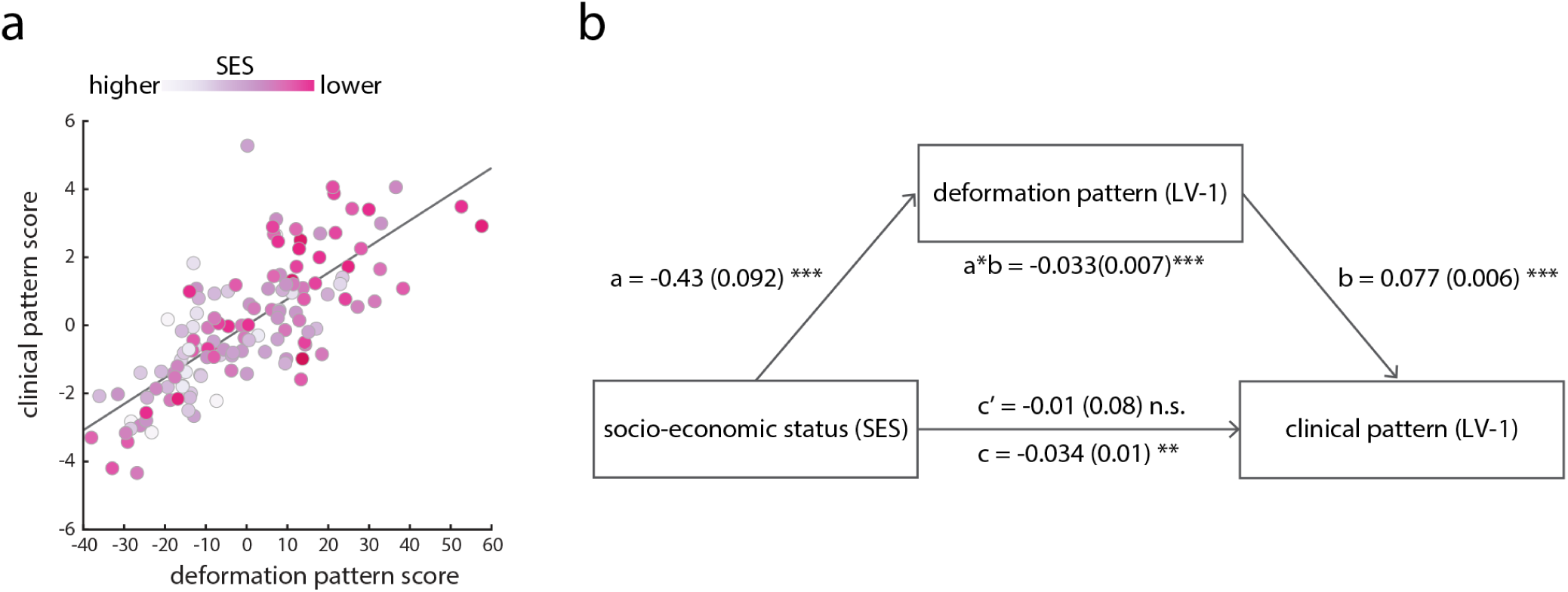
Mediation analysis. (a) Correlations between patient-specific scores on the deformation and clinical-cognitive patterns in LV-1 (shown previously in Fig. 2d). Individual points (representing individual patients) are colored by their socioeconomic status (SES); individuals with lower SES tend to score more highly on both patterns. (b) Mediation analysis testing the hypothesis that the effect of SES on clinical-cognitive outcome is mediated by neuroanatomical changes. Regressing the PLS-derived brain deformation pattern on SES showed that lower SES is significantly associated with decreased brain volume (*a* = −0.43(0.092); *P <* 1.0 *×* 10^−4^ ; 95% CI [−0.62, −0.24]. Regressing clinical expression (LV-1) on the brain deformation pattern (LV-1) and SES showed a significant effect of brain deformation on clinical expression (*b* = 0.077(0.06); *P <* 1.0 *×* 10^−4^ ; 95% CI [0.065, 0.089]). However, the direct effect of SES on clinical expression (*c* = −0.034(0.01); *P <* 1.0 *×* 10^−2^ ; 95% CI [−0.05, −0.01]) did not remain significant after the deformation pattern was modeled as a mediator (*c*′ = −0.001(0.008); *P* = 0.9; 95% CI [−0.016, 0.014]). In contrast, the LV-1 brain deformation pattern significantly mediates the effect of SES on clinical expression [*a ∗ b* = −0.033(0.007); *P <* 1.0 *×* 10^−4^ ; 95% CI [−0.049, −0.018].

We next used mediation analysis (details in *Supplementary Results* to ask whether the brain deformation pattern of LV-1 mediates the effect of SES on the corresponding clinical outcome (symptom severity) (Fig. 3b). Unstandardized parameter estimates and standard error for the model are shown in (Fig. 3b). Regressing the PLS-derived brain deformation pattern on SES showed that lower SES is significantly associated with decreased brain volume (*a*) (Fig. 3b). Regressing clinical expression (LV-1) on the brain deformation pattern (LV-1) showed a significant effect of brain deformation on clinical expression (*b*). However, the direct effect of SES on clinical expression (*c*) did not remain significant after the deformation pattern was modeled as a mediator (*c*^*t*^) (Fig. 3b). In contrast, the LV-1 brain deformation pattern significantly mediates the effect of SES on clinical expression (*a * b*) (Fig. 3b). Taken together, the mediation analysis reveals an indirect-only mediation (mediated effect *a * b*) with brain deformation as mediator. In other words, severity of brain abnormalities mediates the effect of lower SES on clinical expression of the cognitive-negative dimension.

As socioeconomic status is often confounded with race, we stratified patients into Caucasian and African-American and directly compared their clinical and deformation scores. Fig. S4a suggests that African-American patients tend to have greater LV-1 clinical and deformation scores, but that this is mainly explained by differences in socioeconomic status (two-sample *t*-test:*t*(131) = 3.70, *P* = 3.15× 10^−*4*^; significantly lower socioeconomic status for African-American patients). Critically, the relationship between brain deformation and clinical scores can be observed in each group separately (Fig. S4a, *r* = 0.76, *P* = 5.03 × 10^−*13*^ and *r* = 0.69, *P* =4.00 × 10^−*11*^ for Caucasian and African-American patients, respectively). Moreover, the two correlation coefficients were not significantly different (Fisher’s test, *Z* = 0.83, *P* = 0.41), suggesting that the first clinical-anatomical dimension remains a viable measure across different races.

## DISCUSSION

In the present report we used multivariate mapping of comprehensive clinical features and voxel-wise brain deformation to isolate latent clinical-anatomical dimensions of schizophrenia. Three latent clinical-anatomical dimensions were identified, collectively accounting for 55% of brain-behavior covariance, but only the first (27%) was replicated in an independent dataset. This clinical-anatomical dimension encompassed cognitive deficits and negative symptoms, and mapped onto a distributed brain deformation pattern centered on the default mode and visual networks. Brain deformation of this cognitive-negative dimension was represented across different races but was more pronounced in patients with lower SES. These findings suggest that a considerable population variance in schizophrenia can be described by a compact set of continuous multimodal phenotypic axes, mainly shaped by cognitive-negative symptoms and network-specific anatomical abnormalities.

### Multimodal heterogeneity of schizophrenia

Understanding the heterogeneity of clinical and anatomical manifestations of schizophrenia remains a major challenge in schizophrenia research^31^. Numerous studies have investigated single symptom domains in relation to either global measures (e.g. total brain volume, global cortical thickness)^15,18,19^ or localized brain abnormalities in pre-defined regions of interest^17,20^. And yet, symptom domains in schizophrenia often occur simultaneously (e.g. secondary negative symptoms due to positive symptoms and/or depression,^72,73^) and are highly correlated (e.g. cognitive deficits and negative symptoms,^74–76^). Likewise, brain abnormalities covary across structurally and functionally connected regions^22,23^. In the present report, we take a step towards a more comprehensive and multimodal understanding of the disease. Using a single integrative analysis, we find that the complex constellation of clinical-behavioral and anatomical features can be parsimoniously summarized by a smaller set of latent clinical-anatomical dimensions. In doing so, we derive continuous, multimodal markers of individual disease status that can be easily computed in new patients and datasets, and are readily comparable with other continuous or categorical solutions.

Importantly, the present results complement modern efforts to derive transdiagnostic biotypes. For instance, Clementz and colleagues used comprehensive neurocognitive and neurophysiological data to identify three discrete biotypes across the schizophrenia spectrum (bipolar, schizoaffective and schizophrenia)^34^. A subsequent voxel-based morphometry study showed that biotype 1 with poor cognitive-sensory function had a broadly distributed cortical and subcortical volume reduction, while biotype 2 with moderate cognitive impairments exhibited more regional volume reduction within the insula and fronto-temporal regions^77^. Transdiagnostic symptom dimensions have been identified in the same dataset, with more severe negative symptoms for biotypes 1 and2 ^78^. Our findings enrich insights from this work, showing a dimensional clinical (cognitive-negative) and neuroanatomical pattern that effectively bridges biotypes 1 and 2. Consistent with Reininghaus and colleagues^78^, the present study demonstrates successful integration of phenomenological and neuroimaging data to identify dimensional characteristics of schizophrenia. Altogether, the identified clinical-anatomical dimension can be readily applied in concert with categorical biotypes, to advance progress in treatment development and diagnostics across the schizophrenia spectrum^34,35,77^.

### Default mode and visual networks - anchors of the cognitive-negative dimension

In the present model, the dominant cognitive-negative dimension was most closely related to deformation in the default mode and visual networks. Our group and others have recently demonstrated that deformation topography in schizophrenia reflects anatomical and functional network topology, with core deficits observed in the default mode network^13,22,23^. For instance, Wannan and colleagues observed a similar network-based pattern of brain abnormalities across multiple stages of the schizophrenia spectrum (from first episode to chronic and treatment-resistant patients)^22^. The present findings extend this work by showing that network based deformation can be mapped to a cognitive-negative dimension.

Previous case-control studies revealed that clinical subtypes with predominantly negative symptoms^79^ and biotypes with cognitive-sensory impairments^77^ demonstrated most extensive cortical thinning and global grey matter reduction respectively. At the same time localized associations have been reported for cognitive function and volume reduction in the anterior cingulate, insula, hippocampus/parahippocampal gyrus, middle frontal gyrus and cognitive function^18,19^ as well as negative symptoms and reduced orbitofrontal cortical thickness^17^. The present report builds on these findings, demonstrating that a cognitive-negative dimension of schizophrenia reflects targeted abnormalities in spatially-specific networks.

More generally, these results shed new light on how pathological processes of brain structure and function are intertwined^10,21,80^. An emerging literature points to a continuous unimodal-transmodal cortical synaptic hierarchy^81,82^, manifesting as smooth topographic gradients of gene transcription^83,84^, intracortical myelin^85,86^, cortical thickness^87^, excitation-inhibition balance^88^, and macroscale structural and functional connectivity^63,89–91^. Our results show that the dominant cognitive-negative dimension originates from the ends or “anchors” of this hierarchy: the visual^15,92^ and default mode networks^93–95^. This raises the possibility that multiple pathological processes, originating from opposing ends of the putative sensory-fugal hierarchy, may be involved in the disease. Interestingly, two recent functional imaging studies found evidence of atypical functional connectivity and integration between unimodal and transmodal cortices^96,97^. Our results show that these deficits in functional coordination may ultimately originate from underlying anatomical abnormalities, reflecting large-scale molecular and cellular gradients.

### Limitations and future directions

The present report highlights a latent clinical-anatomical dimension of schizophrenia, but the findings should be interpreted with respect to several important limitations. First, data-driven multivariate models seek to map multiple modalities to one another, but as a result, they have cannot be used to make inferences about localized relationships between specific clinical symptoms and specific brain regions. Second, the present findings are based on cross-sectional data, precluding extrapolation of longitudinal progression. In addition, there is consistent evidence for five specific psychosis domains of positive, negative, disorganized, manic and depressive symptoms across the schizophrenia spectrum^78^. The current study was based on datasets assessing psychotic symptoms with the SAPS and SANS, which limited the ability to investigate clinical-anatomical dimensions in the presence of additional measures of the affective domain (mania, depression) and a more comprehensive disorganization domain. Future work should employ multivariate approaches including all five symptom domains.

In terms of methodology, it is important to note that head motion could systematically bias structural MRI^98–101^. Addressing this potential confound would require additional in-scanner head-motion estimates from fMRI^99,101^, which were not available in either dataset. Finally, the influence of drug exposure on brain structure is another important confounding factor that is challenging to address in cross-sectional studies. We found no evidence of an association between current medication dose and the clinical-anatomical dimensions in a sub-sample (n=87) of the discovery dataset. However, these results are limited by the fact that current medication does not allow conclusions to be drawn on long-term drug exposure. Future studies in longitudinal data are warranted to explore medication effects on latent clinical-anatomical dimensions.

## Conclusion

The present work contributes to a growing recognition that individual clinical symptoms do not occur in isolation, nor can they be precisely mapped to a single locus in complex disorders such as schizophrenia. An integrated multivariate model allows clinical experience and objective neuroanatomical measurements to simultaneously inform one another, yielding a more holistric understanding of heterogeneity in the patient population. The clinical-anatomical dimension identified here opens a new direction for dimensional stratification, complementing existing efforts to develop sensitive diagnostics and individualized treatment strategies.

## Data Availability

Deformation patterns of the clinical-anatomical dimensions are freely available for visualization and download for further exploration and future research from Neurovault (https://identifiers.org/neurovault.collection:6825).

## Acknowledgments

This research was undertaken thanks in part to funding from the Canada First Research Excellence Fund, awarded to McGill University for the Healthy Brains for Healthy Lives initiative. MK acknowledges support from the National Bank Fellowship (McGill University) and the Swiss National Foundation (P2SKP3_178175). BM acknowledges support from the Natural Sciences and Engineering Research Council of Canada (NSERC Discovery Grant RGPIN #017-04265) and from the Fonds de recherche du Québec - Santé (Chercheur Boursier). GS acknowledges support from the Natural Sciences and Engineering Research Council of Canada (NSERC) and from the Healthy Brains for Healthy Lives (HBHL) initiative at McGill University. CM acknowledges support from the Canadian Institutes of Health Research (CIHR). Data collection and sharing for this project was funded by NIMH grant 1R01 MH084803.

## Data Availability Statement

Deformation Patterns of the clinical-anatomical dimensions are freely available for visualization and download for further exploration and future research from Neurovault (https://identifiers.org/neurovault.collection:6825).

## Supplementary Methods and Results

### Methods

#### NUSDAST inclusion criteria

The initial sample submitted to MRI pre-processing comprised a total of 267 participants (153 patients, 114 controls). Eight participants were removed after failing MRI processing quality control (see *Quality control* below) resulting in data from 146 individuals with schizophrenia (52 female, 34.7±12.9 years) and 113 healthy controls for further analyses. The behavioral (clinical and cognitive) measures as well as demographics and other general information (i.e., medication information, age of disease onset, years of education, socioeconomic status,etc) were also obtained for the individuals with schizophrenia. Among the participants with schizophrenia, 10 individuals were removed due to missing behavioral data (*>* 20% missing values) and 3 individuals were removed due to uneven race distribution relative to the rest of cohort (removed races were Asian-Pacific Islander (*n* = 1) and Hispanic (*n* = 2); the rest were either African-American (*n* = 71) or Caucasian (*n* = 62); for more details see *Clinical and cognitive measures*). Altogether, the final dataset comprised 133 individuals with schizophrenia and 113 healthy controls (selection flowchart is shown in Fig. S1). Data collection and sharing was approved by the local institutional review board and was funded by NIMH grant 1R01MH084803^38^. Informed consent was obtained from each participant after a complete description of the study was given^18^.

#### NUSDAST sample selection and imputation

The NUSDAST database provides a large battery of behavioral data (demographics, clinical and cognitive assessments) and neuroimaging data. We collected a subset of the behavioral data for the individuals with schizophrenia from their baseline visit for our purposes. It should be noted that some of the individuals with schizophrenia were missing parts of the behavioral data. To address this issue, we first removed the individuals who were missing more than 20% of the behavioral variables. Second, we imputed the missing values for the remaining *n* = 133 individuals using K Nearest Neighbors imputation (MATLAB function: knnimput.m). The number of neighbors was set to *k* = 15 (≈ 10% of the total number of individuals) and Euclidean distance was set as the distance metric, such that the missing entry of a variable was replaced by the median value of that variable from its 15 nearest neighbors.

#### Douglas inclusion criteria

Participants inclusion criteria, MRI acquisition and processing were derived from the previous publication from Beland and colleagues^56^. Individuals (n=143) meeting diagnostic criteria for schizophrenia or schizoaffective disorder for a duration of at least 3 years, and aged between 18 and 50 years old, were recruited from inpatient and outpatient units of the Douglas Mental Health University Institute and affiliated community centers. Participants were recruited as a part of a larger cross-sectional study investigating the determinants of insight in schizophrenia. Of this group, 114 patients accepted to participate in the neuroimaging part of the study. Information on diagnosis, antipsychotic dosage (converted to chlorpromazine equivalent), and duration of illness were collected by medical chart review, or directly confirmed with patients’ medical teams. An abbreviated version of the Structured Clinical Interview for DSM-IV Axis I Disorders was administered to all patients to confirm patients’ illness history. Exclusion criteria included low neuropsychological performance, lifetime or familial history of neurological condition, head injury with loss of consciousness, diagnosis of substance dependence, and presence of metallic objects in the body. Additionally, 71 healthy controls, without any personal or familial history of psychotic illness were recruited using a classified advertising website in Montreal. The Structured Clinical Interview for DSM-IV-TR Axis 1 Disorders, non-patient version (SCID-NP) was administered to all healthy controls during the first assessment to rule out the presence of current mental illness. Healthy controls were recruited based on their education level, age, and sex, to match the demographic characteristics of the patient group. All participants provided written informed consent, and the study procedures were approved by the Douglas Mental Health University Institute human ethics review board.

#### MRI acquisition parameters

##### NUSDAST dataset

For details of T1 MRI data acquisition see^38^. All MR scans were collected using the same 1.5 T Vision scanner platform (Siemens Medical Systems). Acquisition of all scans was performed at the Mallinckrodt Institute of Radiology at Washington University School of Medicine, where scanner stability (e.g., frequency, receiver gain, transmitter voltage, SNR) and artifacts were regularly monitored. Whole brain 3-dimensional T1-weighted magnetization prepared rapid acquisition gradient (MPRAGE) scans with the following were applied. Acquisition parameters were: TR = 9.7 ms, TE = 4 ms, flip angle = 10°, ACQ = 1, 256 × 256 matrix, 1× 1 mm in-plane resolution, 128 slices, slice thickness 1.25 mm, 5:36 min per scan.

##### Douglas dataset

T1-weighted structural images were acquired on a Siemens 3 T Tim trio MRI at the Brain Imaging Centre of the Douglas Mental Health University Institute. The scans were MPRAGE (TR = 2300ms, TE = 2.98ms, FOV 256mm, 1× 1× 1 mm voxels, flip angle =9°) and lasted 9 min.

#### Quality control of pre-processed MRI data

##### NUSDAST dataset

Pre-processing of T1 MRI scans from 267 participants was performed using (1)the `minc-bpipe-library` pipeline (https://github.com/CobraLab/minc-bpipe-library). Manual quality control of images included the following steps of inspection: (1) registration of each individual brain relative to the template, (2) proper coverage of individual brain mask on the brain to avoid over/under segmentation, (3) uniformity of intensity profiles across the brain to ensure good inhomogeneity correction. Participants were excluded if images failed in one of the three steps. In total eight subjects were excluded including seven patients with schizophrenia and one healthy control.

##### Douglas dataset

Pre-processing of T1 MRI scans and manual quality control was performed as outlined above for the NUSDAST discovery dataset. After visual quality control of images six patients with schizophrenia and two healthy controls were excluded leaving with a total sample of 108 individuals with schizophrenia and 69 healthy controls for further analysis.

#### Deformation-Based Morphometry (DBM)

Local change in the brain tissue’s volume density was calculated using Deformation-Based Morphometry (DBM;^55^). Regional DBM values can be considered as measures of tissue loss or tissue expansion^39–43^. DBM is estimated based on the deformation applied at each voxel to non-linearly register each brain to a given template.

In the present study, we used the ANTs Multivariate Template Construction pipeline^44^ (`antsMultivariateTemplateConstruction2.sh`) to measure the DBM values. This pipeline produces a population average through the iterative estimation and application of affine non-linear warps to a starting rigid model, in this case the *MNI ICBM 09c symm* model. The final iteration of non-linear transformation of each structural brain image to the unbiased template image produced during the registration process is used as a deformation map for each subject in the template space. A deformation map quantifies the displacement of each voxel in each direction in the 3-dimensional template space that was required to transform a participant’s image to the template. Local change in tissue density is then estimated as the derivative of the displacement of a given voxel in each direction. The derivatives can be calculated as the determinant of the Jacobian matrix of displacement, **J**, which is given as^43^ :

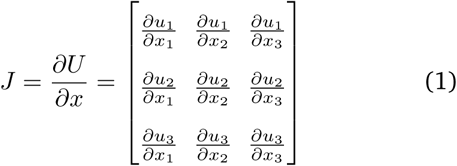

where *x* = (*x*_1_, *x*_2_, *x*_3_) is a position in a given T1-weighted image at the subject space, which is then transformed to the template space in each direction using the displacement given by the vectors **U** = (**u**_**1**_, **u**_**2**_, **u**_**3**_). Each element of the Jacobian matrix is estimated using a first order approximation. As an example, *J*_13_ can be calculated as following:

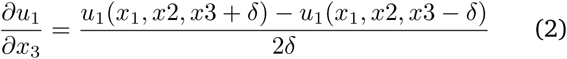

where *δ* is the position of a given voxel along axis *x*_3_. Local changes in tissue density are given by the determinant of the Jacobian matrix: no change in volume is given with 1 (i.e., no displacement relative to template), tissue expansion is given with a value between 0 and 1 (i.e., subject image was shrunk to be transformed to template space), and tissue loss is given with a value larger than 1 (i.e., subject image was expanded to be transformed to template space). For more intuitive interpretation of the changes in volume density, we calculated the logarithm of the determinant, such that no change is given by 0, tissue loss is given by a positive value, and tissue expansion is given by a negative value.

Finally, the deformation maps were blurred (2 mm full-width/half-maximum) and the non-brain tissue was removed from each brain using FreeSurfer image analysis suit (release v6.0.0; documented and freely available for download at http://surfer.nmr.mgh.harvard.edu/). The voxel-wise data was then extracted for each participant for further analysis (see *Statistical model*).

### Clinical and demographic measures for post-hoc analysis with PLS results

Socioeconomic status (SES) was left out from the PLS analysis in order to run post-hoc analysis of the relation between clinical-anatomical dimensions and SES. In addition, age of onset and duration of illness were also preserved for post-hoc analysis, because (a) both variables significantly correlated with age, which was already regressed out from the DBM data (age of onset:rs=0.25, *P* = 3.6 ×10^−3^; duration of illness: rs=0.78, *P* = 1.3 ×10^−28^) (b) we aimed to directly correlate those measures with the clinical-anatomical dimensions to investigate a potential relation between the latent dimensions and disease course. Finally, chlorpromazine equivalents were tested for associate with clinical-anatomical dimensions post hoc.

### Statistical model

Partial least squares is a form of reduced rank regression that identifies linear combinations of two sets of variables that maximally covary with each other. In the present study, one set represents the voxel-wise deformation for each schizophrenia patient (denoted as **X**_*n*×*p*_), while the other set corresponds to the behavioral measures (denoted as **Y**_*n* × *q*_). The *n* rows of both matrices **X** and **Y** represent the number of schizophrenia individuals (i.e., *n* = 133). The *p* columns of matrix **X** correspond to the number of voxels. Matrix **X** was first thresholded above zero and only the positive values corresponding to volume loss were retained. The DBM values in matrix **X** were then corrected for the age of schizophrenia patients using a linear regression model. The *q* columns of matrix **Y** correspond to the behavioral measures from the same schizophrenia patients. We included 15 demographic and clincial measures in the PLS analysis (See *Clinical and cognitive measures*). Finally, both **X** and **Y** matrices were standardized column-wise (i.e., z-scored) and a correlation matrix (**X***′* **Y**) was computed from the standardized matrices. Singular value decomposition (SVD)^103^ was then applied to the correlation matrix **R** = **X**′ **Y** as follows:

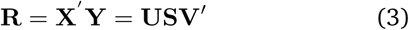

The decomposition results in two orthonormal matrices of left and right singular vectors (**U** and **V**, respectively), and a diagonal matrix of singular values (**S**). The main results of PLS analysis are represented as latent variables, which are constructed from the 3 resulting matrices (i.e., **U, V**, and **S**). More specifically, latent variables are mutually orthogonal, weighted linear combinations of the original variables of the two initial data blocks (i.e., **X** and **Y**) and express the shared information between the blocks with maximum covariance. Latent variable *i* (LV_*i*_) is composed of the *i*th column vector of **U**, *i*th column vector of **V**, and the *i*th singular value from **S**. The elements of the column vectors of **U** and **V** are the weights of the original voxel-wise deformation values and behavioral measures, respectively, that contribute to the latent variable. The weighted deformation and behavioral patterns maximally covary with each other, and the covariance between them is reflected in the corresponding singular values from the diagonal elements of matrix **S**. The effect size associated with the latent variable *i* can also be measured using the singular values, as follows:

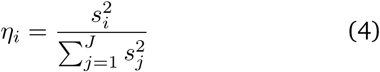

where *η*_*i*_ is the effect size for LV_*i*_, *s*_*i*_ is the corresponding singular value from the diagonal matrix **S**, and *J* is the total number of singular values. Furthermore, the PLS-derived deformation and behavioral patterns (i.e., left and right singular vectors, **U** and **V**) can be used to estimate patient-specific scores that demonstrate the extent to which each patient expresses the patterns. The patient-specific deformation and behavioral scores are calculated by projecting the PLS-derived deformation and behavioral patterns (i.e., **U** and **V**) onto the original patient data:

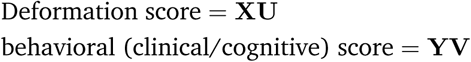

To assess the model, we performed four additional steps: (a) statistical significance of overall patterns was assessed by permutation tests^57^ ; (b) feature (voxel, clinical measure) importance was assessed by bootstrap resampling^58^ ; (c) out-of-sample correlations between projected scores were assessed by cross-validation^59^ ; (d) stability of deformation and clinical patterns was assessed by split-half resampling^60^. We discuss each below. *Permutation tests*. We assessed the statistical significance of each latent variable using permutation tests^57^. During each permutation, the rows of data matrix **X** were reordered randomly and a new permuted correlation matrix was calculated using **Y** and permuted **X**. Similar to the original analysis, the permuted correlation matrix was then subjected to SVD. The procedure was repeated 10,000 times resulting in a null distribution of singular values. To test the null hypothesis that there is no specific relationship between deformation values and behavioral measures, a *P*-value was estimated for each latent variable as the proportion of the times that the permuted singular values were greater than or equal to the original singular value.

#### Bootstrap resampling

The reliability of singular vector weights (i.e., weights of voxel-wise deformation values and behavioral variables) were assessed using bootstrap resampling (10,000 repetitions)^58^. The rows of data matrices (i.e., **X** and **Y**) were randomly resampled with replacement and new correlation matrices were calculated using the resampled data matrices. The correlation matrices were then subjected to SVD as before, generating a sampling distribution for each deformation and behavioral weight in the singular vectors. To assess the reliability of each variable, we calculated bootstrap ratios as the ratio of each variable’s weight to its bootstrap-estimated standard error. Bootstrap ratios allow us to identify variables (voxels or clinical measures) that make a large contribution to the overall pattern (i.e. have a large weight) and, at the same time, are stable across individuals (i.e. have a small standard error). If the bootstrap distribution is Gaussian, a bootstrap ratio can be interpreted as a z-score^58^, such that 95% and 99% confidence intervals correspond to bootstrap ratios of ± 1.96 and ± 2.58, respectively.

#### Cross-validation

We used cross-validation to assess the out-of-sample correlation between deformation and clinical scores^59,104^. We used 100 randomized train-test splits of the original data, where 75% of the data was treated as a training set and 25% of the data was treated as an out-of-sample test set. For each training set, PLS was used to estimate deformation and behavioral patterns (i.e., **U**_*train*_ and **V**_*train*_). Next, the test data were projected onto the deformation and behavioral patterns derived from the training set. This allowed us to estimate patient-specific scores and their correlation for the test sample (i.e. *corr*(**X**_*test*_**U**_*train*_, **Y**_*test*_**V**_*train*_)). This procedure was repeated 100 times to generate a distribution of out-of-sample correlation coefficients. Finally, we used permutation tests (100 repetitions) to assess the significance of these out-of-sample correlation coefficients. During each permutation, we randomly shuffled rows of the original deformation matrix **X** and repeated the above procedure. The procedure generated a null distribution of correlation coefficients between deformation and clinical scores in the test sample. This null distribution was then used to estimate a *P*-value, by calculating the proportion of correlation coefficients that were greater than or equal to the mean original out-of-sample correlation coefficient.

#### Split-half resampling

Finally, we sought to assess the stability of PLS patterns using split-half resampling^60^. The original data were randomly split into two halves and new correlation matrices were calculated for each half separately (i.e., **R**_**1**_ and **R**_**2**_). Singular value decomposition was performed for one half as follows:

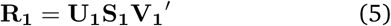

The correlation matrix from the other half was then projected onto the singular vectors derived from the first half (i.e., **U**_**1**_ and **V**_**1**_) to calculate the new singular vectors for the second half (i.e., **U**_**2**_ and **V**_**2**_):

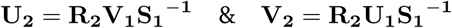

The procedure was repeated 100 times. For each split, we correlated the projected singular vectors of one half with the ones from the other half (i.e., **U**_**1**_ and **U**_**2**_, and **V**_**1**_ and **V**_**2**_). The final stability measure is the mean correlation coefficient across splits.

## RESULTS

### Reliability of clinical and deformation patterns

Although Fig. 2d depicts strong correlations between clinical scores and deformation, the analysis is designed to maximize these values, increasing the possibility of overfitting the statistical model. To assess the statistical reliability of the patterns, we perform two additional analyses. First, we assess the out-of-sample correlation between clinical and deformation patterns^104^. We repeated the analysis with 100 randomized train and test splits, representing 75% and 25% of the sample, respectively. Test data were projected onto the PLS models estimated from the training set. The predicted clinical and brain deformation scores were then correlated (Fig. 2e). As expected, the mean out-of-sample correlations were lower than the in-sample correlations (LV-1: *r* = 0.27; LV-2: *r* = 0.14; LV-3: *r* = 0.16), but were statistically significant against a permutation test (100 repetitions) with the exception of LV-2 (LV-1: permuted *P <* 0.01; LV-2: permuted *P* = 0.06; LV-3: permuted *P* = 0.02).

Second, we assess the stability of the clinical and deformation patterns by split-half resampling^60^ (see *Supplementary Methods*). Briefly, the sample is split into halves and singular value decomposition is performed for one half. Data from one split are then projected onto the singular vectors (corresponding to clinical and deformation patterns) derived from the other split. Pattern stability is then estimated as the mean correlation between the singular vector calculated directly from one split and the singular vector estimated by cross-projecting data from the other split. The mean correlations among projected clinical and deformation patterns were as follows: LV-1: *r* = 0.69 (95% CI: [0.64 0.73]) and *r* = 0.09 (95% CI: [0.09 0.10]); LV-2: *r* = 0.20 (95% CI: [0.15 0.25]) and *r* = 0.06 (95% CI: [0.05 0.08]); LV-3: *r* = 0.19 (95% CI: [0.14 0.25]) and *r* = 0.04 (95% CI: [0.03 0.06]). Consistent with previous reports, the clinical patterns were more stable across splits compared to the brain deformation patterns^28,59^.

### Clinical and anatomical features of LV-2 and LV-3

Fig. S2b shows the loadings (i.e. correlations) of individual, clinical and cognitive scales with the second latent variable (LV-2). Please note that LV-2 is represented by negative loadings on all significant measures, indicating lower cognitive performance and also lower symptom scores. Reduced cognitive functioning (WAIS-Voc, *r* =. −39, 95% CI [−.25,.53]; WMS-Cog, *r* = −.21, 95% CI [−.02,-.36]; WAIS-Matrix, r=-.2, 95% CI [−.07,-.4]),educational attainment (*r* =. −26, 95% CI [−.11,-.42]), low positive symptoms (SAPS Disorganization Global, *r* =. −29, 95% CI [−.22,-.47]; SAPS Disorganization Sum, *r* = .28, 95% CI [−.21,-.47]) low diminished emotional expression symptoms (SANS Diminished Expression Global, *r* =. −31, 95% CI [−.24,-.5]; Diminished Expression Sum, *r* =. −33, 95% CI [−.25,-.53]) and lower educational attainment (*r* = −.26, 95% CI [−.11,-.42]) were the strongest contributors to LV-2. Low levels of Avolition-Apathy (SANS Avolition-Apathy Domain) and DIMD contributed also significantly to LV-2 but to a lesser extent (all *r ≤*.2). In other words, the LV-2 captures a clinical phenotype of cognitive deficits in the relative absence of positive and negative symptoms (cognitive-only dimension).

Fig. S2b (left panel) shows the corresponding deformation pattern, indexed by bootstrap ratios (see *Materials and Methods*). This deformation pattern is comprised of parietal, dorsolateral prefrontal (superior frontal gyrus, middle frontal gyrus, inferior frontal gyrus and anterior cingulate, as well as subcortical regions including the striatum and brainstem. Altogether, the pairing indicates that across patients, greater deformation in these regions is associated with severity of cognitive deficits but not with positive or negative symptoms. To further demonstrate the relation between the clinical measures and anatomical maps, we projected individual patient data onto the PLS-derived left and right singular vectors to estimate patient-specific scores. Fig. S2c shows the correlation between the clinical and deformation scores (*r* = 0.68).

Fig. S2c shows the loadings (i.e. correlations) of individual, clinical and cognitive scales with the third latent variable (LV-3). The strongest contributors to LV-3were: gender (*r* = −.58, 95% CI [−.45,-.67]) (greater deformation in males), higher positive symptoms (SAPS Disorganization Global, *r* = .30, 95% CI [.16,.52]; SAPS Disorganization Sum, *r* = .19, 95% CI [.08,.40]); (SAPS RealityDistortion Global, *r* = .21, 95% CI [.06,.40]), low/diminished emotional expression symptoms (Diminished Expression Sum, *r* = −.37, 95% CI [−.20,-.49]; SANS Diminished Expression Global, *r* = − .28, 95% CI [−.13,-.44]), higher Avolition-Apathy scores (SANS Avolition-Apathy Global, *r* = .23, 95% CI [.06,.42];SANS Avolition-Apathy Sum, *r* = .20, 95% CI [.03,.39]), lower educational attainment (*r* =. −22, 95% CI [−.04,-.37]) but good cognitive performance (WAIS-Matrix, *r* = .2, 95% CI [.06,.42]). In other words, LV-3 captures a clinical-anatomical dimension particular prevalent in male patients (gender-related dimension) with combined positive symptoms and motivational deficits (Avolition-Apathy score).

Fig. S2c (left panel) shows the corresponding deformation pattern, indexed by bootstrap ratios (see *Materials and Methods*). This pattern is comprised of lateral occipital (cuneus), parietal (supramarginal gyrus) and frontal regions (precentral gyrus, superior, middle and inferior frontal gyri). Altogether, the pairing of this third clinical-anatomical dimension indicates that across patients, greater deformation in these regions is more prevalent in male patients and more strongly associated with positive symptoms and motivational deficits. To further demonstrate the relation between the clinical measures and anatomical maps, we projected individual patient data onto the PLS-derived left and right singular vectors to estimate patient-specific scores. Fig. S2c shows the correlation between the clinical and deformation scores (*r* = 0.82).

### Mapping of LV-2 and LV-3 on intrinsic networks

LV-2 and LV-3 were mapped on intrinsic networks and subjected to the same spatial permutation procedure used for the main analysis in LV-1^62^. The deformation pattern of LV-2 (cognitive-only dimension) was significantly associated with the frontoparietal and ventral attention networks (*P* = 8.4 × 10^−*3*^ and *P* = 1.9 × 10^−*2*^, respectively; (Fig. S2b). The deformation pattern of LV-3 (gender-related dimension) was significantly associated with the dorsal attention network (*P* = 2.2 × 10^−*3*^; (Fig. S2c). Altogether, these results demonstrate that the identified clinical-anatomical dimensions are linked to organized deformation patterns, centered to specific intrinsic networks.

### Effects of age of onset, duration of illness and medication dosage

In an exploratory analysis, we assessed whether patient-specific clinical and brain deformation scores are related with age of onset, duration of illness or medication dosage. We found a statistically significant association between age of onset and brain deformation as well as clinical scores of the LV-1 and LV-3 (LV-1, clinical: *rs* = 0.19, *P* = 0.03, brain: *rs* = 0.22, *P* = 0.01; LV-3, clinical: *rs* = 0.25, *P* = 0.004, brain: *rs* = 0.28, *P* = but not with clinical scores of LV-2 (LV-2, clinical: *rs* = 0.05, *P* = 0.59, brain *rs* = 0.17, *P* = 0.05). In contrast, duration of illness was not associated with clinical scores or brain deformation (LV-1, clinical: *r* = −0.02, *P* = 0.85, brain: *r* = − 0.05, *P* = 0.32; LV-2, clinical: *r* = −0.10, *P* = 0.59, brain: *r* = −0.03, *P* = 0.56),LV-3, clinical: *r* = −0.06, *P* = 0.26, brain *r* = 0.15, *P* = 0.09.) Information on current medication dosage (chlorpromazine equivalents) were available for a subset of 87 individuals with schizophrenia. We found no statistically significant association between medication dosage and clinical scores or corresponding deformation patterns of all three LV (LV-1, clinical: *r* = −0.02, *P* = 0.88, brain: *r* = −0.10, *P* = 0.32; LV-2, clinical: *r* = 0.07, *P* = 0.51, brain: *r* = −0.03, *P* = 0.76), LV-3, clinical: *r* = −0.13, *P* = 0.21, brain *r*− = 0.22, *P* = 0.19).

**Figure S1.**
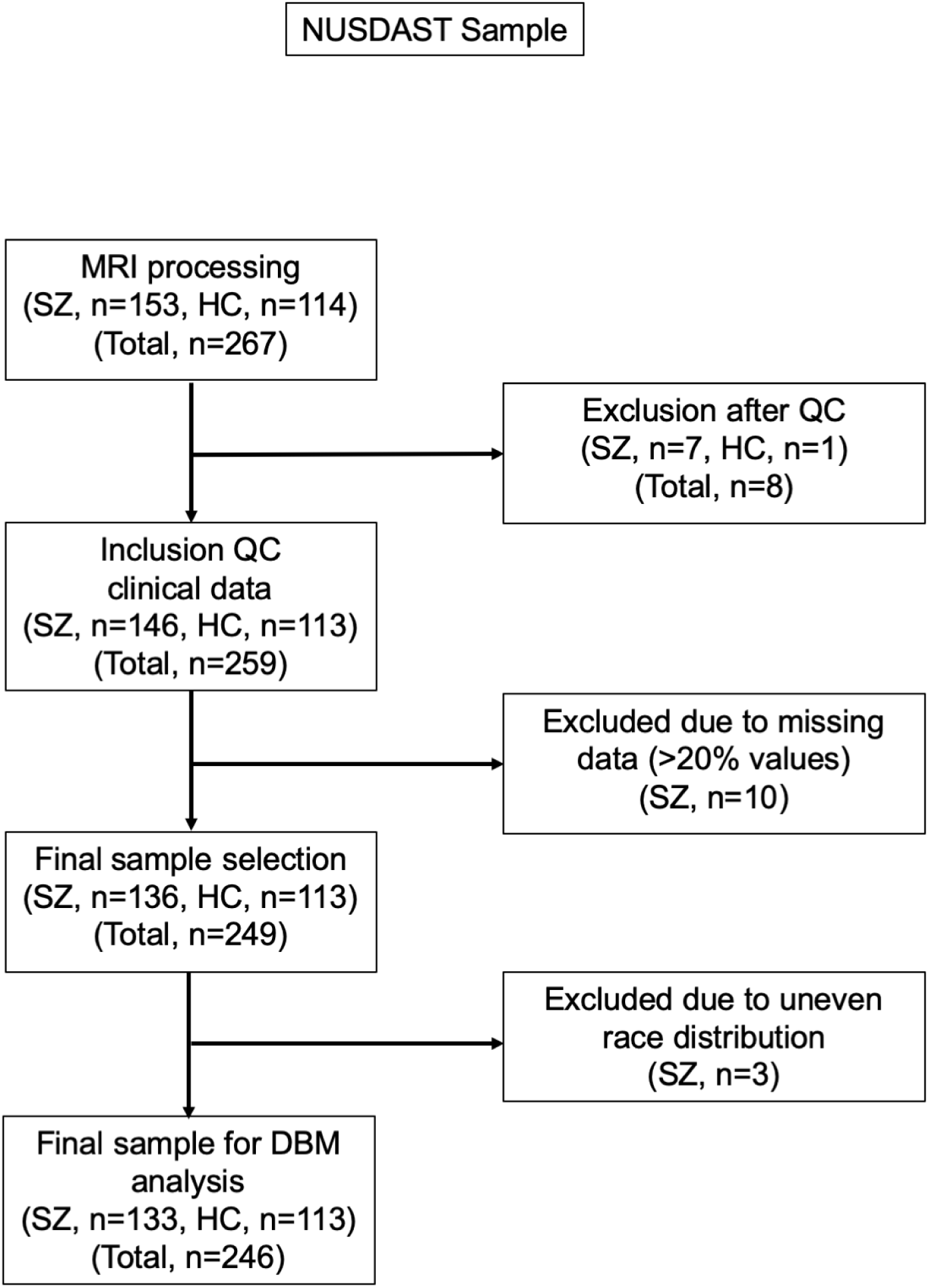
Flow chart of NUSDAST sample selection.

**Figure S2.**
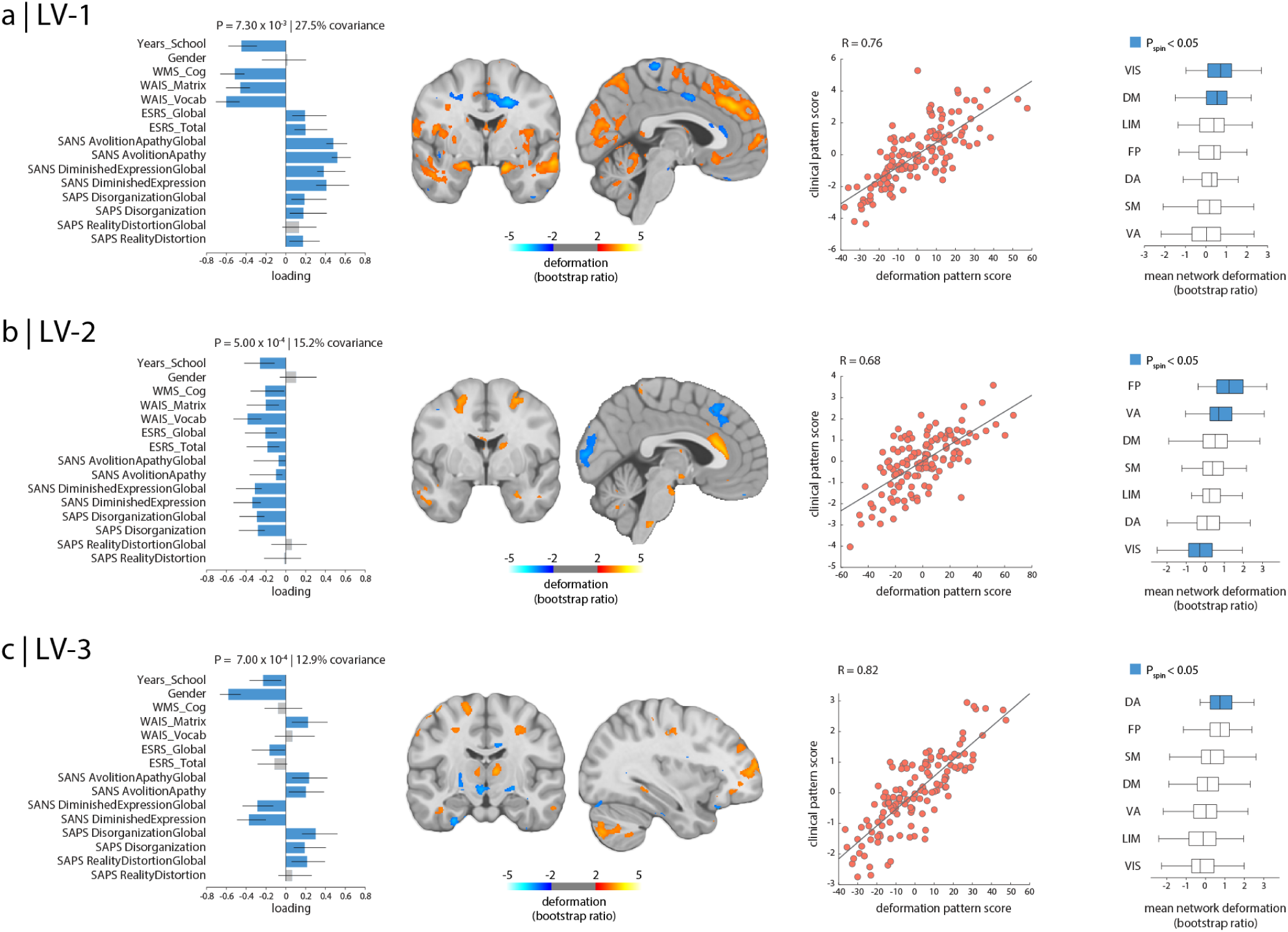
Relating deformation and clinical manifestation. Partial least-squares analysis detected three statistically significant latent variables; LV-1 (a), LV-2 (b) and LV-3 (c), mapping distributed patterns of deformation to clinical-behavioral phenotypes. First column: LV clinical features. The contribution of individual clinical measures is shown using correlations between patient-specific clinical scores and scores on the multivariate pattern (loadings). Error bars indicate bootstrap-estimated standard errors. Second column: LV deformation pattern displayed on an MNI template (MNI152_symm_2009a; LV-1: *x* = − 3, *y* = − 2; LV-2: *x* = −3, *y* = − 1; LV-3: *x* = −34, *y* = −14). The contribution of individual voxels is shown using bootstrap ratios (ratios between voxel weights and bootstrap-estimated standard errors; see *Materials and Methods* for more detail). Patients who display this deformation pattern tend to score higher on positively-weighted clinical measures and lower on negatively-weighted clinical measures. Third column: individual patient data is projected onto the weighted patterns shown in the first and second columns to estimate scalar patient scores that quantify the extent to which individual patients express each pattern in the LV. Fourth column: system-specific deformation. The PLS-derived deformation pattern is stratified into resting-state networks (RSNs) defined by Yeo and colleagues^61^. The bars indicate mean deformations for each network. *P*-values are estimated with respect to the spin test null developed by Alexander-Bloch and colleagues^62^. Yeo networks: DM = default mode, DA = dorsal attention, VIS = visual, SM = somatomotor, LIM = limbic, VA = ventral attention, FP = fronto-parietal.

**Figure S3.**
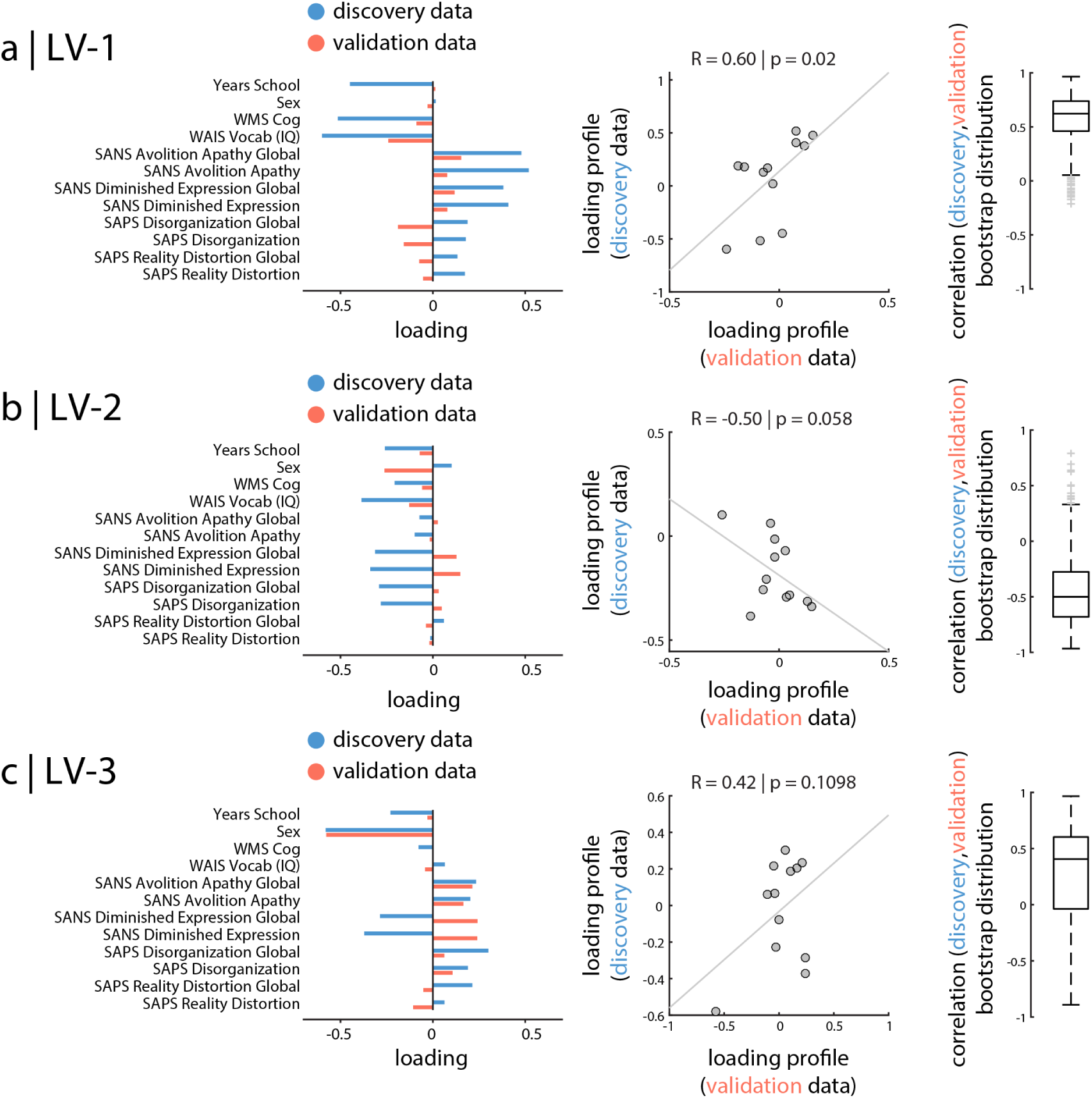
Replication dataset. The statistical PLS models derived from the discovery dataset (NUSDAST; blue) was applied to the validation dataset (Douglas; orange). Brain imaging data (i.e., DBM maps) from the validation set were projected onto the latent variables derived in the discovery set (Fig. S2), yielding a predicted clinical profile. The two datasets were compared on a reduced set of 12 overlapping clinical, cognitive and demographic measures. Note that IQ and total cognitive scores are estimated differently in the two datasets. Vocabulary subtest of Wechsler Adult Intelligence Scale (WMS-III;^54^) was used as a measure of crystallized knowledge (i.e., premorbid crystallized intellectual functioning (ePMC-IQ)^18^) in the discovery dataset, while Weschler Abbreviated Scale of Intelligence (WASI full-scale IQ;^56^) was used in the validation dataset. A composite score of overall cognitive functioning was estimated from subtests of the Wechsler Memory Scale (WMS-III;^54^) in the discovery set, whereas the composite cognitive score for validation dataset was estimated from the CogState Research Battery protocol^102^ that includes cognitive domains of verbal memory, visual memory, working memory, processing speed, executive function, visual attention, and social cognition. Left column: clinical profile from the discovery data (blue) and projected clinical profile from the validation data (orange). Data are shown as loadings. Middle column: scatter plots between the disocvery and validation clinical profiles. Pearson correlations and permuted *p*-values are shown. Right column: bootstrap-estimated distributions of the correlations between discovery and validation profiles are depicted.

**Figure S4.**
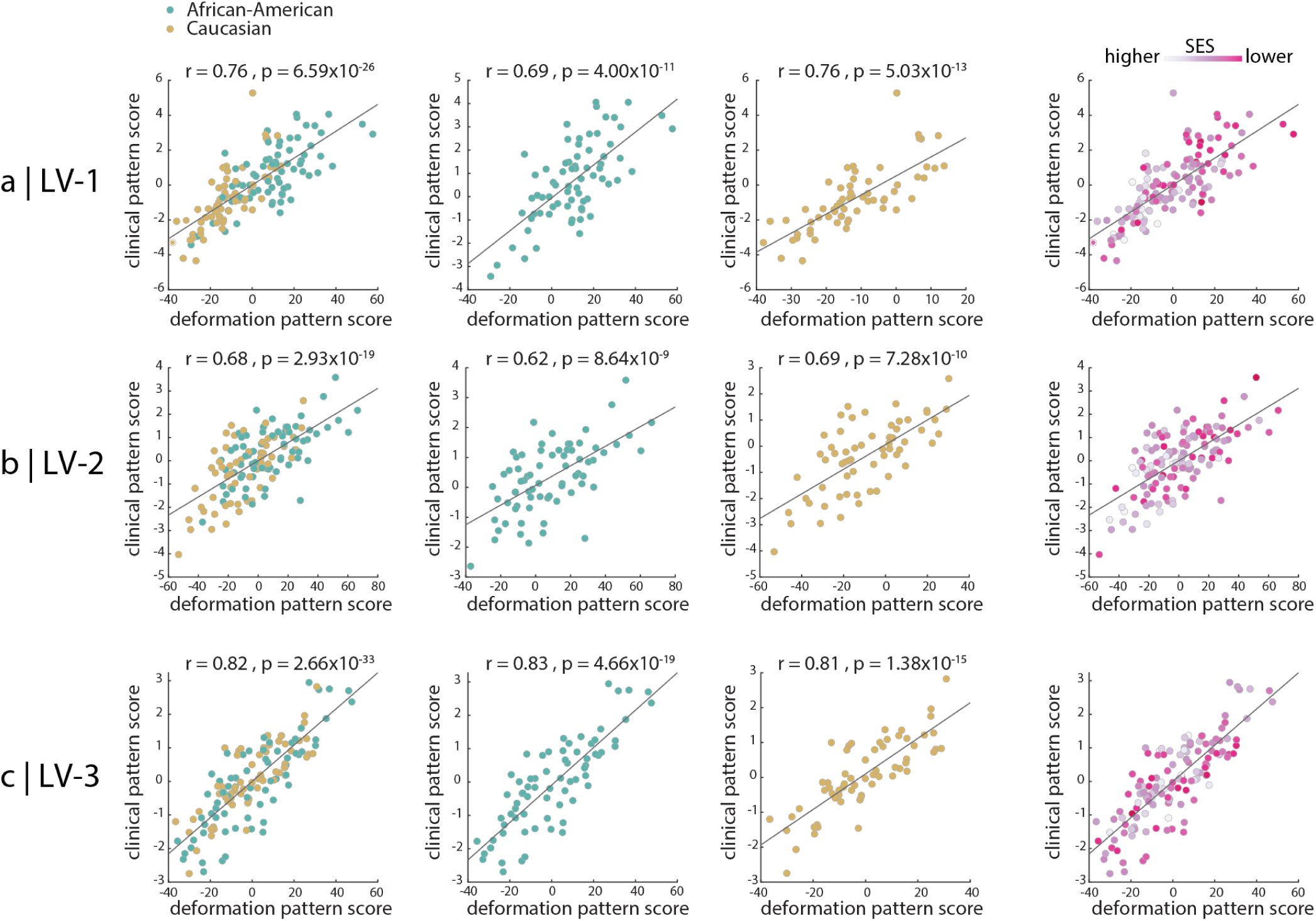
The effect of race and socio-economic status on clinical and anatomical patterns. Participants are stratified by race for the three latent variables (panels a-c). First column: linear models, relating clinical and anatomical patterns, are fit for African-American and Caucasian participants jointly. Second and third column: linear models, relating clinical and anatomical patterns, are fit for African-American and Caucasian participants separately. Fourth column: patient-specific clinical and anatomical scores are colored by the individual’s socio-economic status.

